# Impact of Electronic Immunization Registries and Electronic Logistics Management Information Systems in four Low-and Middle-Income Countries: Guinea, Honduras, Rwanda, and Tanzania

**DOI:** 10.1101/2025.01.27.25321171

**Authors:** C Mantel, C Hugo, C Federici, N Sano, S Camara, E Rodriguez, L Castillo, J Condo, P Irakiza, I Sabi, E Nyanda, W Olomi, M Cavazza, V Mangiaterra, M Verykiou, E Ferenchick, A Torbica, T Cherian, S Malvolti

## Abstract

**Background:** There is increasing interest in low-and middle-income countries (LMICs) to introduce and scale-up digital health tools like electronic immunization registries (eIR), and electronic logistics management information systems (eLMIS) to support immunization services. An evaluation of the use of these tools was conducted in four countries to inform decisions about their further expansion and investments.

**Methods:** Purposive sampling of regions, districts, and health facilities was done in each country based on predefined criteria. Primary data were collected between October 2021 and September 2022 in 50 health facilities in Guinea, 88 in Honduras, 36 in Rwanda, and 101 in Tanzania using semi-structured questionnaires, standardized competency assessments and data accuracy checks. Data focused on electronic tool usage, user experience, infrastructure, workforce needs, and decision-making, as well as immunization data quality and perceptions of health workers and vaccine recipients. Data analysis combined both quantitative and qualitative methods.

**Findings:** The implementation of eIR and eLMIS was associated with improvements in National Immunization Programme (NIP) processes and outcomes. Users were satisfied with the tools (87% satisfaction rate), and 95% of users in Africa valued the accessibility of information, with 91% finding it accurate and complete. Some caregivers reported better organization and shorter waiting times in health facilities using the tools. Most eIR users noted improvements in process efficiencies (81%) and immunization service delivery (89%). In Rwanda and Tanzania data accuracy was higher in exclusively paper or electronic settings (60%) compared to dual paper-electronic systems (45%). eLMIS use was associated with improvements in vaccine stock data quality and reduced stock-outs. While 77% of health workers were digitally literate, inadequate digital infrastructure was a key barrier to tool use. Interoperability with the Civil Registration and Vital Statistics system (CRVS) was limited, hindering the tracking of unimmunized children.

**Conclusions:** To fully realize the potential of electronic tools in LMICs, full government ownership, targeted infrastructure investments, migration to fully electronic systems, and integration of eIR with the CRVS are essential.

## Introduction

Immunization plays a critical role in preventing disease and protecting lives and is considered one of the most successful and cost-effective public health interventions. The World Health Organization (WHO) estimates that vaccination currently averts between 3.5 to 5 million deaths every year (1). The COVID-19 pandemic highlighted the significant impact of vaccination on outbreak control, while widespread disruptions in health systems resulted in setbacks in routine immunization. In the attempt to restore immunization coverage and to further strengthen immunization programmes, accurate information is required on both vaccination status at the individual level and on the performance of vaccine supply chains. However, data quality in LMICs is considered inadequate, and will need to be enhanced by improving health workers’ capacities, data collection tools, reporting and feedback (2). High-quality and timely data to inform decision-making are necessary to support immunization service delivery, track under- or unimmunized individuals, prevent vaccine stock-outs, and improve operational planning to increase vaccination coverage and reduce morbidity and mortality from vaccine-preventable diseases.

There is a growing interest from governments, donors and implementing partners to introduce and scale-up digital health interventions to support immunization and maternal and child health services (3–6). Countries are using electronic data technologies to advance their immunization information systems, shifting from traditional paper-based systems to electronically collect, process and analyze information (7–9). Innovative “e-health” tools, including eiR and eLMIS, have been piloted, many of which were also newly developed during the COVID-19 pandemic. eIR are computerized, population-based data systems that contain individual-level information on vaccine doses received (10). eLMIS are systems of technology-based records and reports used to collect, organize, present and use logistics data gathered across all levels of the immunization system (11).

While several of these digital tools had been implemented in higher income countries with varying degrees of success, there is an increasing interest from LMICs to use and evaluate these systems in supporting their immunization systems and services (7, 12–21), coupled with an increasing demand for technical and financial support for their further development and implementation.

Earlier assessments of the implementation and impact of eIR and eLMIS highlighted differences across regions and countries. Data on eIR use was collected using online surveys in Europe, Latin America, and Asia (5, 7, 22). An evaluation in European countries highlighted specific benefits of eIR, including use for linking immunization data with other health outcome and safety data (4). In India, an evaluation of an electronic system to capture individualized information on service delivery for mothers and infants showed a positive impact on data quality and completeness (23). In Tanzania, the deployment of electronic immunization interventions resulted in strengthened data collection, improved data accuracy and completeness and increased use of data for programmatic decision-making (24). Further reviews documented the utility of eIR in monitoring the impact of immunization programmes (16, 25). A study of a combined eIR-eLMIS in Tanzania demonstrated a reduction in vaccine stock-outs by using an electronic rather than a paper-based LMIS (17). An evaluation of an eLMIS in Zambia found that timelines and frequency of stock reporting had increased, and that the system had contributed to improvements in efficiency, cost, and commodity security (26). However, various challenges of implementing such systems across countries were seen, of which infrastructure requirements, interoperability, and data quality were the most widely discussed (27, 28).

## Methods

A multi-country evaluation was conducted with the goal of providing actionable evidence for ministries of health and global financing institutions to inform decisions about the implementation and management of eIR and eLMIS, and future investments in these tools. The evaluation was carried out in Guinea, Honduras, Rwanda, and Tanzania between June 2020 and October 2022 and collected experiences and practical lessons learned on the governance and modalities of the use of eIR and eLMIS. Additional data were collected on the costs, affordability, and sustainability of these systems and an economic impact analysis conducted. Its outcomes are summarized in a parallel publication (29).

The four countries were selected from a comprehensive listing of LMICs with an eIR and/or eLMIS. Countries were shortlisted by applying criteria including income level and geographical location, the duration of use of the tools beyond a pilot phase and the availability of relevant data across health system levels. The final selection was done in a consensus-finding approach in collaboration with senior staff of the WHO, Gavi, the Vaccine Alliance (Gavi) and the Bill and Melinda Gates Foundations (BMGF). The in-country research institutions, Africa Health Consulting in Guinea, the Centre for Impact Innovation and Capacity Building for Health Information and Nutrition (CIIC- HIN) in Rwanda, an independent team of consultants in Honduras, the Mbeya Medical Research Center, National Institute for Medical Research (MMRC-NIMR) in Tanzania, together with the evaluation team of Bocconi University and MMGH Consulting developed the evaluation protocols, and planned and conducted the fieldwork. All protocols and data collection instruments received ethical approval under the procedures set by the Guinea National Health Research Ethics Committee, the Pan American Health Organization (PAHO) Ethics Review Committee (for Honduras), the Rwanda National Ethics Committee, the Tanzania Medical Research Coordinating Committee and the Bocconi University Research Ethics Committee.

Three major research questions guided the programmatic evaluation: i) What is the impact of eIR and/or eLMIS on the NIP and service delivery in terms of process and outcomes? ii) What are the barriers and opportunities for implementing these systems? Iii) How interoperable are the systems with the national HMIS and CRVS?

In each of the four countries, a purposive sampling of regions or provinces, districts, and health facilities was adopted based on predefined criteria including health facility type, size of the catchment population, urban or rural location, Pentavalent (Diphtheria, Tetanus, Pertussis, Hepatitis B and *Hemophilus Influenzae* Type b) vaccine coverage and drop-out rates, status of electronic tool implementation, and time since first use (see Annex 1 for sampling strategies). Primary data were collected in October/November 2021 in Tanzania, in February/March 2022 in Rwanda, in April 2022 in Guinea and in September 2022 in Honduras. A total of 50 health facilities in 4 regions and 7 districts were included in the sample in Guinea, 88 health facilities in 8 health regions in Honduras, 36 health facilities in 5 regions and 12 districts in Rwanda and 101 health facilities in 10 regions and 30 districts in Tanzania. The number of interviews and other data collection efforts are shown in Table 1.

**Table 1.** Number and type of data collection tools completed in the four countries.

|  | Guinea | Honduras | Rwanda | Tanzania |
| --- | --- | --- | --- | --- |
| **Programmatic interviews** |  |  |  |  |
| Health facility | 43 | 80 | 24 | 61 |
| District | 7 |  | 12 | 30 |
| Region |  | 8 |  | 10 |
| **Economic interviews** |  |  |  |  |
| Health facility | 43 | 80 | 24 | 61 |
| District | 7 |  | 12 | 30 |
| Region |  | 8 |  | 10 |
| **Competency assessments** |  |  |  |  |
| Health facility | 43 | 80 | 49 | 25 |
| District |  |  | 16 |  |
| Region |  |  |  |  |
| **On-site accuracy checks** |  |  |  |  |
| Health facility | 43 | 80 | 24 | 62 |
| District | 7 |  |  |  |
| Region |  |  |  |  |
| **Health workers surveys** |  |  |  |  |
| Health facility | 43 | 80 | 44 | 60 |
| **Caregiver interviews** |  |  |  |  |
| Health facility |  |  | 95 | 81 |
| **Total** | **236** | **416** | **300** | **430** |

Data collection instruments were adapted from pre-existing and validated tools, including the Modular Data Quality Assessment Protocol with Electronic Immunization Registry Component (30), data instruments used in the Evaluation of the Better Immunization Data Initiative (31), and the eIR Readiness Assessment Tool*^1^*. Standard interview guides and questionnaires were established and pre-tested in each country in health facilities not included in the sample. Data were collected using portable electronic devices with Open Data Kit (ODK) software and uploaded on central servers via the Kobo Collect application*^2^*. Information was gathered on the use of the electronic tools, the relevant health service infrastructure, workforce requirements including training and supervision, and the use of data for decision-making. Users’ technical experience with the tools was verified via a standardized competency assessment exploring e.g., the addition of new health records, the generation of reports, and their correct interpretation (31). Immunization data quality, accuracy, and timeliness were evaluated by performing selected on-site accuracy checks across different data sources. In addition, perceptions of health workers and vaccine recipients or their caregivers on the usefulness and impact of the tools on immunization performance were elucidated through individual interviews and focus group discussions. Administrative immunization coverage data were obtained from the respective Ministries of Health and organizations involved in implementing the digital tools.

A mixed methods approach involving quantitative and qualitative methods was followed for data analysis. Comparisons were made between the situation before and after the introduction of the electronic systems or between those health facilities that used the new tools and those that either did not use them or only used them to a limited extent. Health workers in facilities where the tools were introduced and still in use at the time of data collection were considered ‘users’, while health workers in facilities where the tool was never introduced, its use discontinued, or used only infrequently were considered ‘Low or non-users’. Subgroup analyses were done by comparing tool use between different levels of the health systems and between urban and rural areas.

The effect of the use of the tools was assessed using a set of predefined service delivery indicators, including factors critical for their successful implementation and scale-up (see Theory of Change Indicators in Annex 2). The analysis related these indicators to the design, functionality and user experience of the tools, and their actual implementation in the respective country, embedded in a health ecosystem depicted by governance, human capacity, infrastructure, and financing.

## Findings

### Country Contexts and Ecosystems

In **Guinea** use of the eLMIS started in 2015, during the Ebola epidemic, when it facilitated the management of vital health commodities. It was thereafter expanded across nine public health programmes, including immunization. The system, based on OpenLMIS v2, was developed locally with support from an external partner. The tool was implemented in a hybrid setup. Data were collected at the health facility level using a paper LMIS version, which was digitized at the district level. As of October 2022, the eLMIS was available in all regional and district health directorates and all 37 public hospitals, as well as in more than half of higher-level health centers. In **Honduras,** the eIR was introduced in 2011, with a pilot phase in two health regions, introduction in additional regions in 2013 and reaching national scale in 2019. By design, the eIR was a dual paper and electronic system. The absence of information technology (IT) hardware in most health centers restricted the use of the digital component to the regional level and to mid-level health facilities, with the remaining health facilities collecting information on paper, subsequently digitized at higher levels. The eIR was not integrated with the eLMIS or the CRVS. In **Rwanda,** the eIR was rolled out nationwide from September 2019 to January 2020 in all 505 health facilities that delivered immunization services. The eIR was based on the DHIS2*^3^* e-Tracker and linked to the national identification number as unique identifier, allowing for interoperability with the CRVS. At the health facility level, health workers either entered data directly into the electronic tool or completed paper-based records which were entered by data managers maintaining and updating the e-Tracker. Data were used for monthly reporting, for generating defaulter lists, new immunization records and for supportive supervision. At the central level, the Rwanda Biomedical Center collated, analyzed, and provided feedback on data obtained from lower administrative levels. In **Tanzania** three electronic tools were in use: an eLMIS, an eIR and a combined platform. The tools were based on customized versions of OpenLMIS*^4^* (for the eLMIS, introduced in 2015) and OpenIZ*^5^* (for the eIR, introduced in 2016). The systems were operational in over 5,000 health facilities in 15 of the 26 regions. The eLMIS was used down to the district level while the eIR was also in use at the health facility level, where it provided the interface for the eLMIS. Some pilot regions had attempted to switch to fully electronic systems, but parallel paper processes were still in place everywhere (32). The eIR was not interoperable with the existing birth registration systems. Table 2 summarizes the tool and implementation status in each of the countries.

**Table 2:** Digital health tools implemented in the four countries. eIR: electronic Immunization Registry; eLMIS: electronic Logistic Management Information System.

| Country (tool) | Guinea (eLMIS) | Honduras (eIR) | Rwanda (eIR) | Tanzania (eIR + eLMIS) |
| --- | --- | --- | --- | --- |
| Local name | eSIGL | SINOVA | e-Tracker | TimR (eIR), VIMS (eLMIS) |
| Technical platform | OpenLMIS | Custom development | DHIS2 eTracker | OpenIZ/SanteSuite (eIR)<br>OpenLMIS (eLMIS) |
| Implementation started | 2018 | 2012 | 2019 | 2014 (eIR); 2015 (eLMIS) |
| Scale at time of evaluation | In 59/444 health facilities | nationwide | nationwide | In 15/26 regions |
| Implementation status at time of evaluation | Parallel paper and electronic process. Paper LMIS used at health facilities, with data back entered in eLMIS available from district level upwards. | No electronic tools at lower-level health facilities. Paper tool data back entered by data clerks or health workers at mid-level primary care centers or regional offices. | Varied use at health facility level alongside paper processes. Data back entered by data clerks and health workers. Not implemented at the regional level. | eLMIS implemented down to the district level; eIR down to the health facility level where it provides the interface for the eLMIS. |

Various models of digital health governance and integration were in use. In both Rwanda and Tanzania, strong political support and strategic frameworks for digital health coupled with experience in employing IT solutions in the health sector aided tool implementation. In Guinea, the introduction of the eLMIS was based on a national strategy for the digitization of logistics and supply chain systems across 14 health programmes. In Honduras, on the other hand, the eIR was rolled out without a strategic digital health framework and with various stakeholders impacting its implementation.

Constraints in access to IT infrastructure affected tool implementation and hindered adoption in urban and rural settings in all countries. While most users charged with data entry had access to hardware (tablets and computers), unreliable electricity and internet connectivity impeded the use of the tools for real-time decision-making (see figure 1). Tool use was further hampered by erratic software updates and poor interoperability between electronic systems.

**Figure 1:**
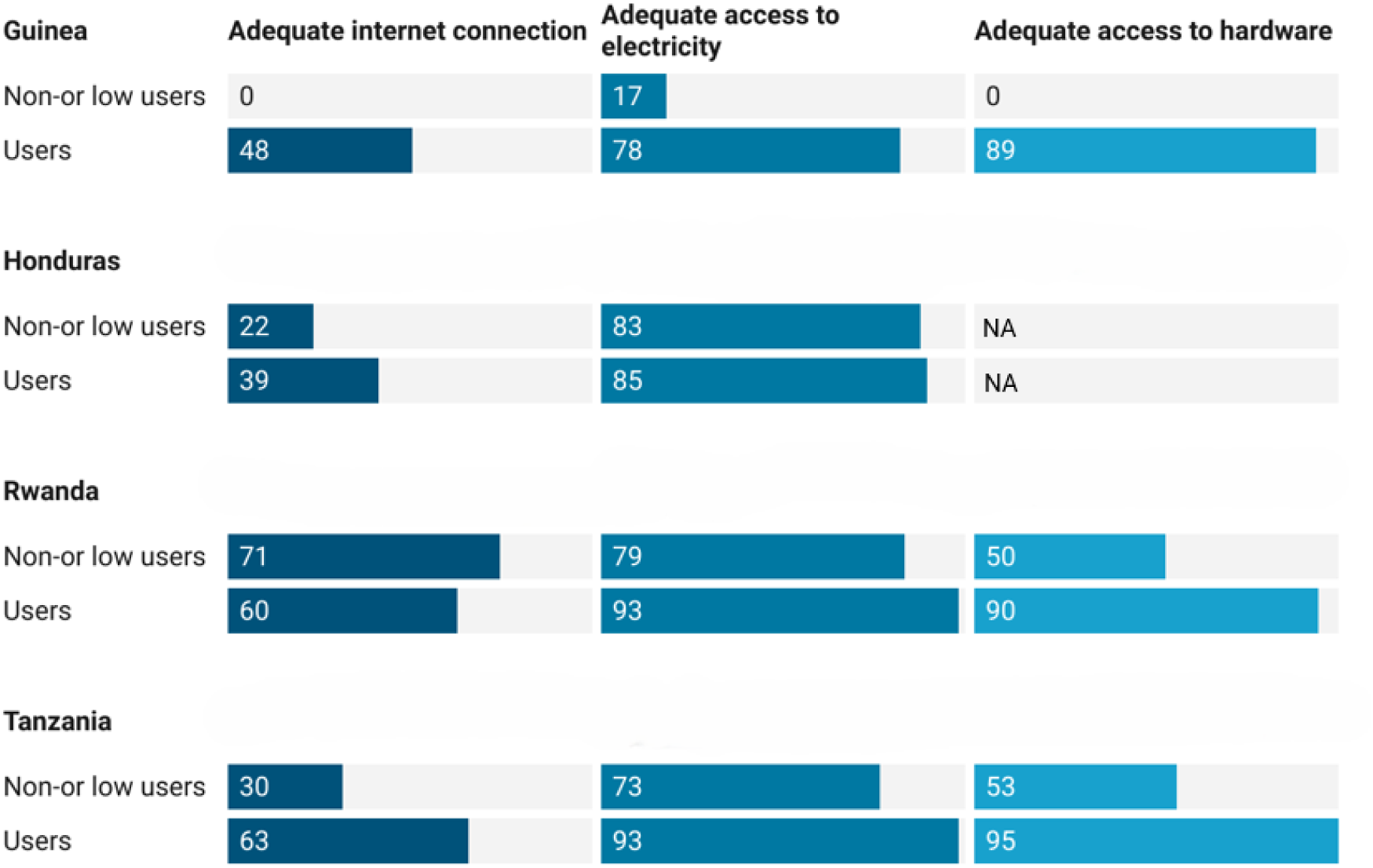
Users’ perception of access to infrastructure (%). Note: In Honduras electronic data entry was not done at the health facility level, hence hardware was not available at this level.

Importantly, digital literacy of health workers was high (81 – 93%) in the three African countries, thus not constituting a barrier to using the electronic systems (see figure 2). eIR users in **Rwanda** and **Tanzania** appeared to be mostly (83% and 87% respectively) competent at completing a new immunization record, as indicated by the standard competency assessments (data not shown). There was limited competence in interpreting the immunization status (41% and 55% respectively) and in generating (21% and 66% respectively) and interpreting defaulter reports (36% and 25% respectively). In **Guinea** , 72% of eLMIS users were able to generate reports on vaccine consumption, while 39% demonstrated advanced skills in accessing stock information.

**Figure 2:**
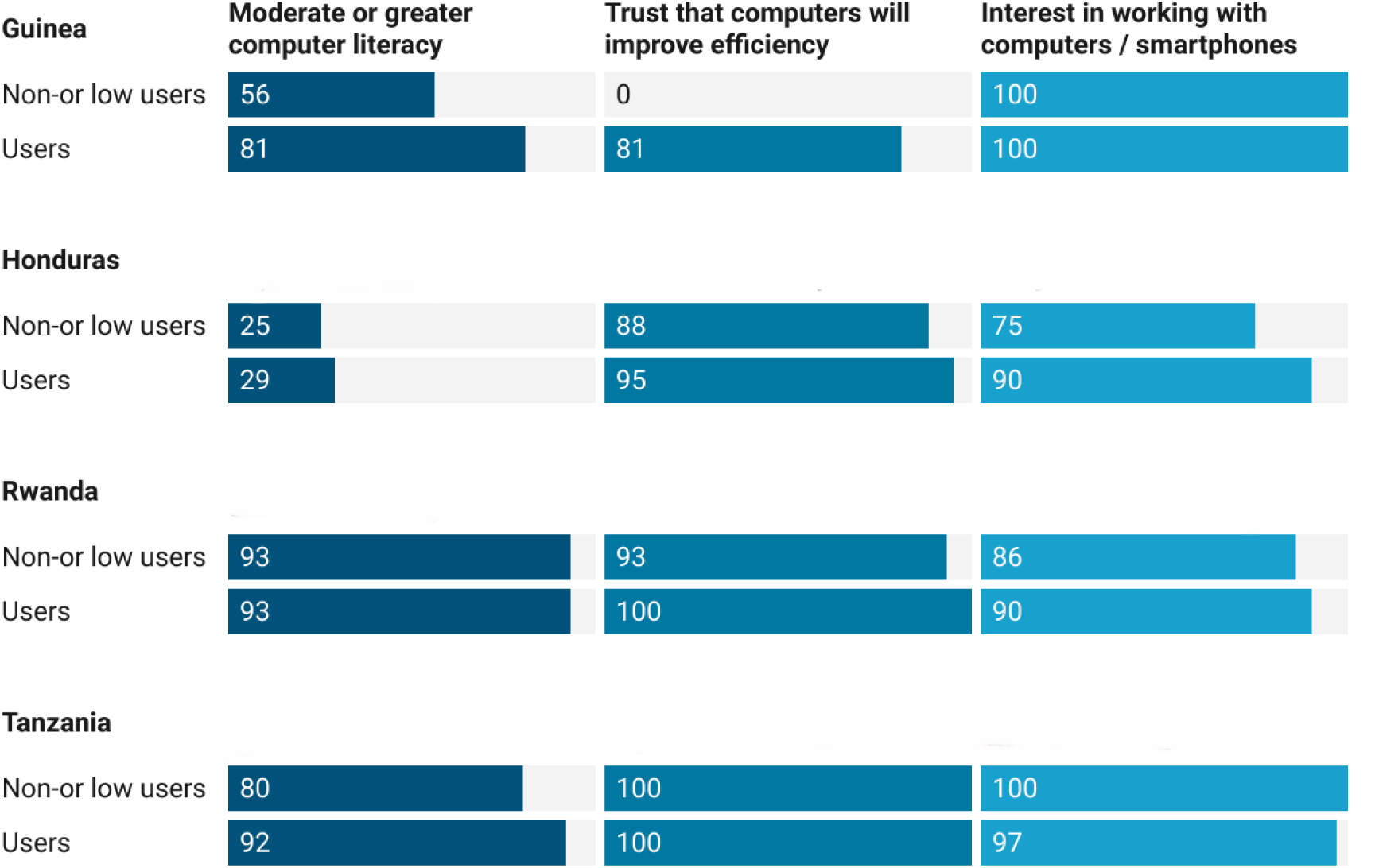
Users’ perception of their own computer literacy and interest in working with electronic tools (%)

### Tool Functionality and Interoperability

Across countries, users were generally satisfied with the quality of IT support, user guides, and supervisor assistance, all factors that were positively associated with tool adoption (figure 3).

**Figure 3:**
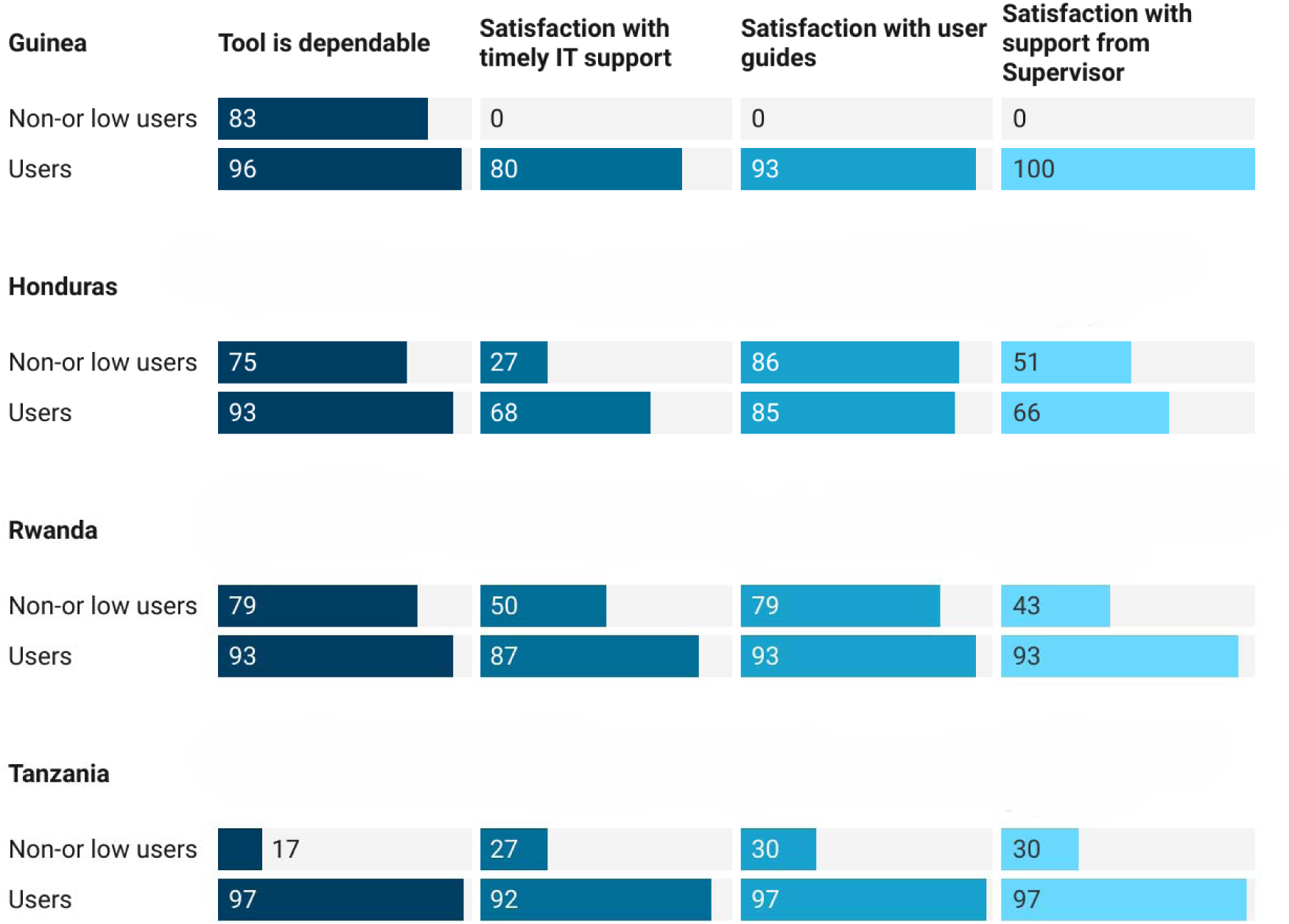
Users’ perception of the quality of tools and services provided (%).

Whilst users in the African countries were highly satisfied with the format of the tools, the accuracy and completeness of data, and their ability to access and use information from the tools, this was only partially true for Honduras (figure 4).

**Figure 4:**
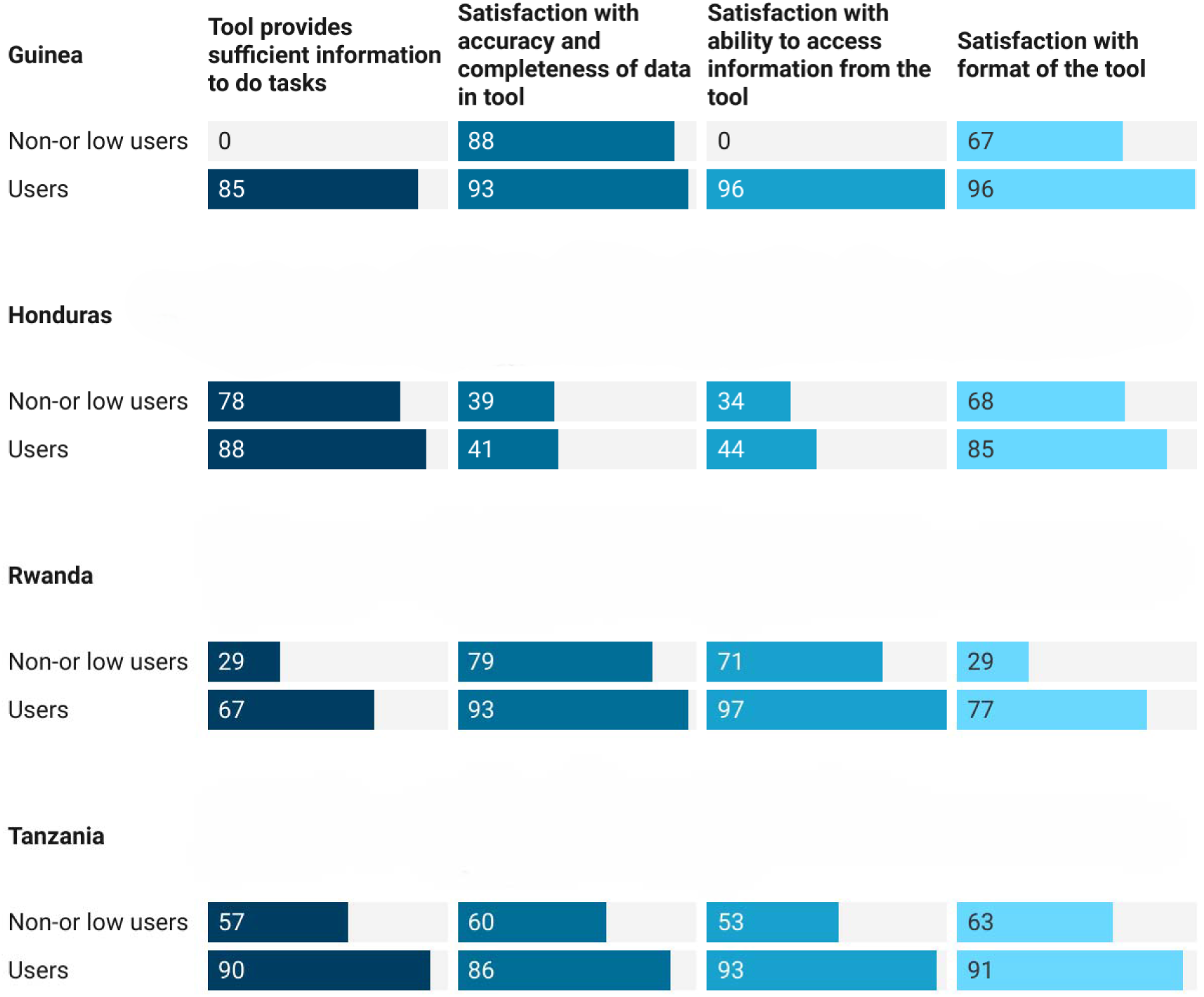
User perception of quality of information in the tool (%)

Policy decisions, design choices, and technical limitations resulted in barriers to the wider use of the electronic tools and in limited interoperability with other health information systems. In **Guinea,** the eLMIS did not contain a working vaccine forecasting or ordering functionality, which impeded its use for stock planning and replenishment at lower levels. While efforts were made to integrate data management platforms across health programs, the eLMIS and the HMIS were interoperable only at the central level and integration approaches led to duplications and specific vaccine programme needs not being met. In **Honduras**, the eIR was set up as a hybrid solution aimed at data validation and reporting at the higher levels. Substantial delays in the registration of newborns limited the feasibility of the integration with the CRVS. In **Rwanda,** the eIR formed part of the National Health Information Exchange System architecture. The COVID-19 pandemic shifted government priorities and delayed the scale-up of the eIR, the use of SMS reminders for vaccine recipients and the transition to full electronic use. Integration with the CRVS was accomplished only in 2024. In Tanzania interoperability between the eIR and eLMIS faced major challenges. To date, the eIR function for sending SMS reminders has only been partly implemented and the tool is not yet interoperable with the CRVS.

### Tool Rollout

In none of the four countries had the electronic tools fully replaced the paper-based legacy systems at the time of the evaluation. In **Guinea,** the eLMIS was not designed to evolve into a fully digital version. In **Honduras**, use of the eIR was limited to the regional level with legacy paper systems remaining in use elsewhere. However, the new immunization registry process allowed for capturing the information necessary for the follow-up of defaulters and for sending reminders. More than two-thirds of the health facility respondents (68%) stated that the new process had led to improvements in data quality and completeness, while 88% indicated that it had positively impacted regional feedback loops. **Rwanda** initiated the nationwide shift to a fully electronic system in October 2022 together with the use of digital vaccination cards and completed this process in early 2024. In **Tanzania,** at the time of the evaluation, the use of the electronic tools had been largely abandoned with one-third of health facilities no longer using them because of unresolved technical challenges. The attempt of some pilot regions to implement fully electronic registry systems remained largely unsuccessful.

External support for maintaining the tools was crucial in all countries. Donors and technical partners drove much of the initial development and implementation processes. However, overreliance on external partners for funding and technical assistance was perceived by some country respondents to distort priorities and generate unwanted dependencies. According to them, deployment of external software developers may also have hindered the acquisition of specialized local knowledge and the ability to adequately respond to technical challenges.

### Tool Use

The electronic tools were inconsistently used across health system levels. At the service delivery level, the tools were most often applied for determining the needs for immunization sessions, for human resources planning and - in Rwanda and Tanzania - for forecasting vaccine requirements. At the district level, EPI supervisors used the tools for programme monitoring and evaluation, monthly reporting, and for adapting supervisory visits. Some supervisors also reported that using the tools had raised the attention to data quality.

Most health workers using the tools were satisfied with their use. Tools were considered trustworthy, user-friendly, and helpful for improving immunization services (see figure 5).

**Figure 5:**
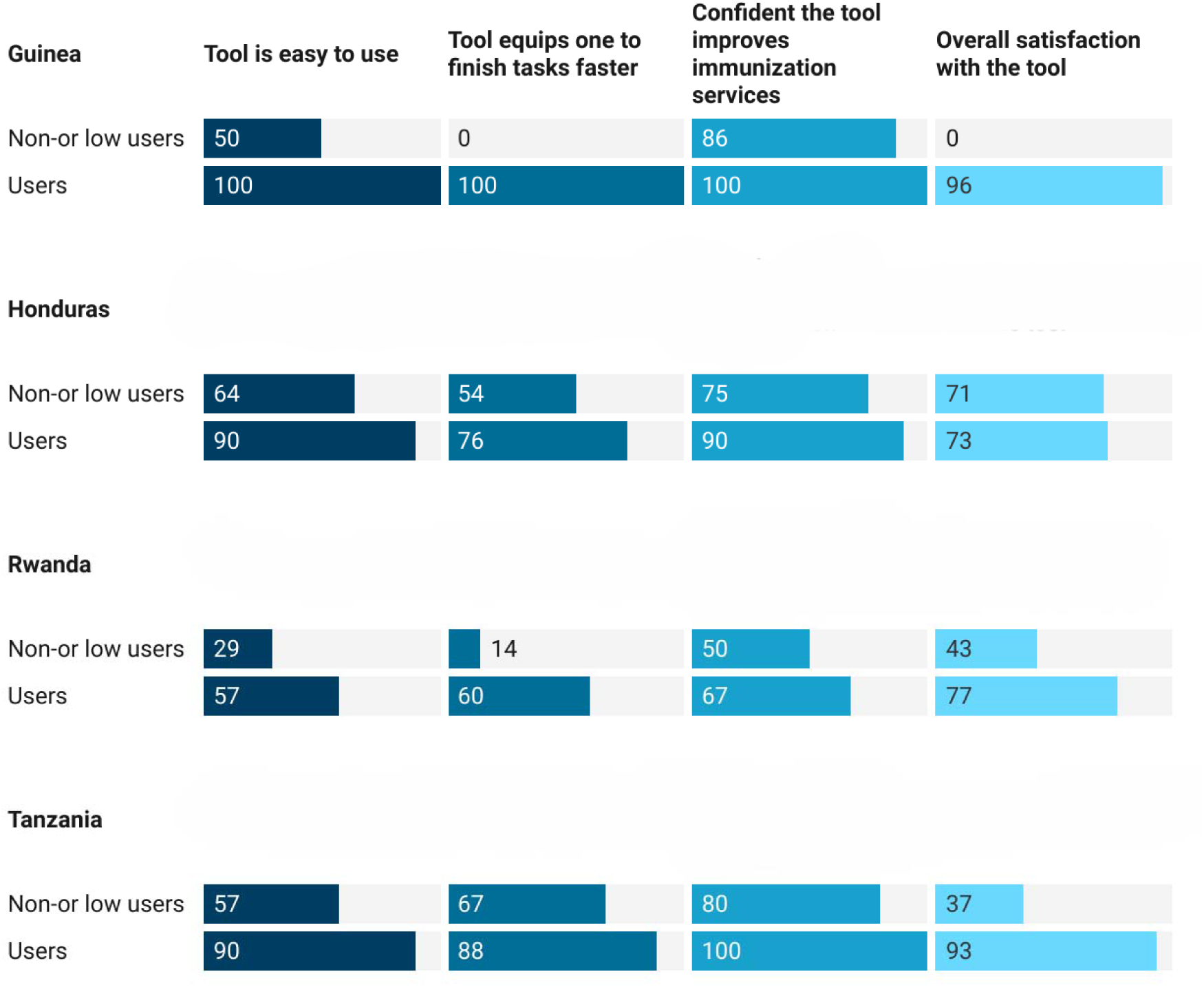
Users’ satisfaction with the tools (%)

Concerns were raised regarding eIR data entry performed by data clerks and not by health workers at the service delivery level or done in batches at certain time points. This practice, often triggered by the absence of hardware in the vaccination room, impeded the ability of health workers to use real-time data for their day-to-day decision-making.

### Tool Impact - eIR

Use of the eIR for tasks in service delivery such as monitoring dropout rates, producing lists of defaulters and reminder messages, generating new home-based records, planning of vaccination sessions, and tracking of supervisory feedback and was highly variable (see figure 6). Differences were mainly driven by the technical capability of the respective tools. Health workers stated that the eIR made it easier to identify and track children beyond the health facility catchment area. Higher level staff indicated that data generated by the tools equipped them to better identify performance gaps.

**Figure 6:**
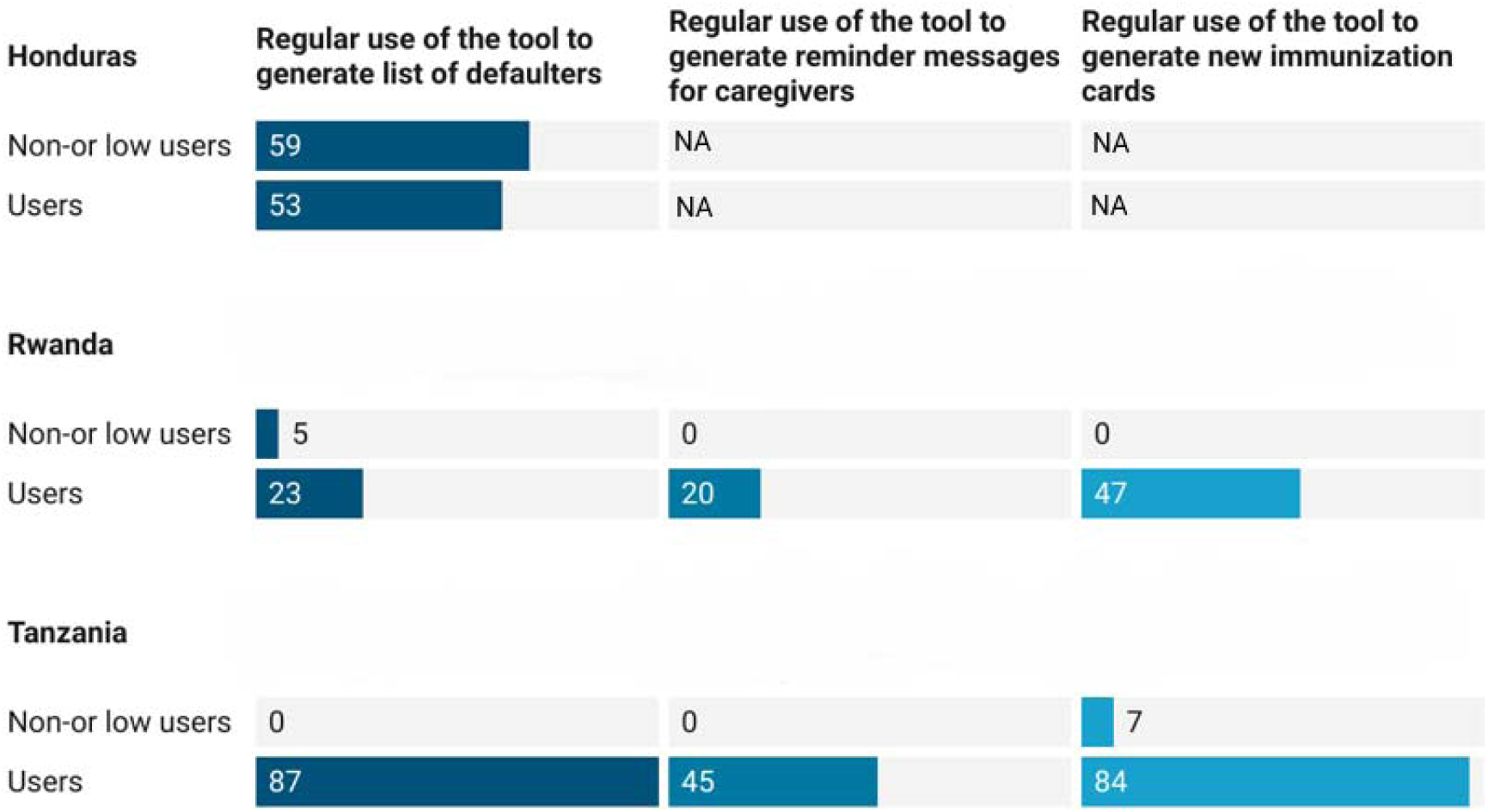
eIR use cases (%). Note: In Honduras there was no electronic data entry at the health facility level. Lists of defaulters were generated at higher levels (with digital tools in place) and communicated to health facilities.

In **Rwanda**, most of the health facility (60%) and district level users (62%) stated that they could finish tasks faster and that their daily work had improved using the eIR. Health facility users, particularly in rural areas, considered the eIR data quality superior to the paper registries. Caregivers in health facilities using the tool found these facilities to be more organized with less waiting times and recognized the prospect of recovering lost home-based records. In **Rwanda and Tanzania** 58% and 83% respectively of health facility and 67% and 90% respectively of district level respondents stated that data quality including accuracy, completeness, and timeliness, and reporting had improved with the introduction of the tools. In Tanzania data quality checks across several variables from three different sources (the electronic registry, the paper register, and the home-based vaccination card) showed higher data accuracy (60%) in exclusively paper or electronic settings than in dual paper-electronic systems (45%). Overall, the continued parallel use of paper and electronic systems reduced potential efficiency gains. Paper systems were still considered the most accurate sources of information of a child’s immunization history in all countries. The accuracy of target population estimates or the identification and tracking of un-immunized children could not be enhanced by using the tools due to the missing linkage with birth registration systems.

### Tool Impact - eLMIS

Users of an eLMIS agreed that tool use had improved vaccine stock data quality and reduced the number of stock-out events. In **Guinea** , the eLMIS served as a reporting and monitoring tool across health programs. Although the tool was not digital at the health facility level, its roll-out added attention to data quality and, according to health workers, led to improved data management practices, supervision and accountability. User satisfaction was high even without the LMIS offering full functional characteristics (11). The majority of eLMIS users in **Tanzania** agreed that vaccine stock data quality had improved with the use of the tool. Health facilities using the tool were less likely to have experienced stock-outs than those using the paper-based system or those without an eLMIS. Data extracted from the eLMIS for the two preceding years also showed that regions using the eLMIS in conjunction with the eIR - i.e. allowing for data entry at the health facility level - experienced fewer stockouts than regions using the eLMIS only at the district level or those using a partially paper-based system (figure 7).

**Figure 7:**
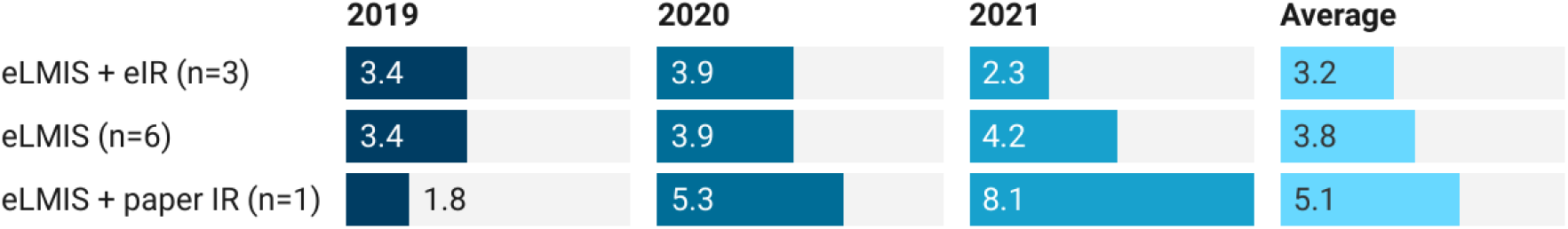
Average number of stock-out events by regional eLMIS use, Tanzania (2019–2021)

### Impact of the COVID-19 Pandemic

The data collection period coincided with the COVID-19 pandemic, which complicated field work across countries. COVID-19 related health system disruptions constrained the deployment and use of the electronic tools with health workers and hardware often being diverted for the pandemic response. Some systems could not reach full national scale or be transitioned to full electronic use or interoperability. At the same time, parallel electronic registration and monitoring systems for COVID- 19 were developed ad-hoc, often only partially aligned with the eLMIS or eIR under evaluation.

Immunization services were negatively impacted by the pandemic. Routine immunization coverage decreased between 2019 and 2021 in all four countries as depicted by Pentavalent vaccine data in the WHO/UNICEF Estimates of National Immunization Coverage *^6^*. Coverage declined from 89% to 81% in Tanzania, from 88% to 77% in Honduras, from 98% to 88% in Rwanda and from 57% to 47% in Guinea. Pandemic-related changes thus prohibited assessing the impact of the use of the electronic tools on immunization outcome indicators such as vaccination coverage, timeliness, or drop-out rates.

## Study limitations

The purposive sampling strategy could have led to sample imbalances resulting in reduced external validity. However, care was taken to select health facilities representative of all those offering immunization based on a-priori criteria. Sample sizes were comparable with those of previous evaluations of eIR and eLMIS implementation in LMICs. Primary data obtained from health workers during surveys and interviews reflected their recollections and perceptions, while there was limited observation of the direct effects of the use of the tools. However, data validation was done by triangulating primary data collected during interviews and focus group discussions with secondary data, e.g., on local service delivery indicators or vaccine stock levels.

## Discussion

Four different tools were evaluated in the four countries, at various levels of implementation: an integrated basic eLMIS in Guinea in the process of being rolled out, a partially electronic IR implemented for several years in Honduras, a newly introduced eIR in Rwanda, and a long-established eLMIS linked with a more recently introduced eIR in Tanzania.

### Tool Ecosystems

Implementing new electronic information systems requires political will, a strategic vision and local expertise. The decision of **Rwanda** to locally build on an existing system (DHIS2) in line with its broader digital strategic vision may have allowed for the ultimately successful implementation of a fully electronic eIR, which, however, at the time of the evaluation, was still being scaled up. In **Tanzania** the eIR, newly customized with extensive external support, was no longer in use by a third of health facilities at the time of the evaluation, due to technical challenges which could not be resolved locally. The switch to a fully electronic system in Tanzania also remained largely unsuccessful, although it had shown a cost-reduction where implemented (29). It is conceivable that overreliance on external support could hamper the sustained implementation of the systems. The hybrid setup of the electronic systems in **Guinea and Honduras** exemplified digitization being implemented at a particular health system level only. In Guinea, the integrated eLMIS was used only at the district level where supply chain decisions about forecasting and reordering were taken and its implementation was found to be cost-neutral. In Honduras, the eIR was limited to the regional level, with paper systems used elsewhere. Its performance was associated with higher costs (29).

In all countries, use of the tools was limited by infrastructure constraints which could often not be resolved in a reasonable time frame. Recent reports from several LMICs found similar operational barriers, including electricity problems, malfunctioning hardware, outdated software and problematic internet connectivity, pointing to the need for investments in data use infrastructure and local skills-building for a full digital transition (14, 33–35).

### Tool Functionality and Interoperability

Health workers were generally satisfied with the electronic tools and considered them user-friendly and reducing their workload. However, different from use in many higher-income countries (36–41), systems were not operational to the extent possible for tracking of defaulters, sending immunization reminders, or forecasting of vaccine demand, mainly due to infrastructure challenges. In Rwanda, Tanzania, and Honduras the immunization tools were only marginally integrated with those of other public health services, while the integration of the eLMIS into the national HMIS in Guinea could well make this system more robust and sustainable. The lack of interoperability of the eIR with birth registration systems continues to impede the identification of un- or under-immunized children and the improvement of target population estimates.

### Tool Rollout and Use

External partners were involved in the implementation of the systems to a varying degree: Guinea, Honduras and Rwanda primarily used domestic resources and local expertise while Tanzania relied on external service providers to resolve emerging IT challenges. Once such support had been phased out, use of the electronic systems was discontinued in several Tanzanian regions. Earlier experience from Tanzania, Zambia and Vietnam had shown that incorporating end user perspectives in the tool design and development were crucial for ensuring ownership and sustainability, while continued financial investments were necessary both for tool development and for its maintenance (19). The sustained adoption of eIR in East African countries was also found to be dependent on adequate health worker staffing, high-quality training, and data-driven supervision (42). While supervision seemed to have benefited from the use of the electronic tools in the four evaluated countries, training on use of the tools was considered unsatisfactory by many health workers, mainly due to the non-availability of on-site training sessions during the COVID-19 pandemic and limited opportunities for re-training. In addition, the lack of digital data entry by health workers at the service delivery level could have prevented ownership and use of data for local decision-making.

The parallel use of electronic and paper-based systems resulted in apparent duplications and inefficiencies. An earlier review of Tanzanian facilities that had temporarily transitioned to full paperless reporting showed that these facilities were more likely to use the system compared to those with parallel reporting systems (43). It is expected that eIR and eLMIS will be used to their full potential once the parallel paper systems have been removed and legacy tools replaced. While attempts to shift to fully electronic use had mostly failed in Tanzania, this shift may ultimately have been successful in Rwanda, albeit later than planned. An updated digital health strategy coupled with extensive local IT expertise might explain some of this difference. The nationwide rollout of the electronic tool in Rwanda with more compressed timelines compared to the multi-year rollouts in smaller pilot projects in Honduras and Tanzania - or earlier in Vietnam and Zambia (19) - could have allowed the system to deliver its benefits early and to positively influence acceptance and adoption.

### Tool Impact

The use of an eIR was considered beneficial by health workers in contexts with appropriate infrastructure. Health workers stated that they were able to finish their tasks faster, and that the tools strengthened the monitoring of vaccination performance, similar to findings from a recent study from Tanzania (44). The potential of eIR for managing defaulters was not fully exploited, but the tools played a key role in the planning of vaccination sessions, the recovery of home-based records, and in the provision of supervisory feedback. Earlier evaluations in Tanzania showed that eIR also allowed for the analysis of service delivery and care-seeking patterns and of risk factors for under-immunization (45). The impact of the COVID-19 pandemic on routine immunization services barred the use of immunization coverage for the assessment of the tools’ impact. Earlier studies in Pakistan, however, were able to show a positive effect on coverage. The Zindagi Mehfooz system used in the Sindh province for registering and tracking individual immunization status was able to generate more accurate population estimates and vaccination targets. Use of the system was associated with an increase in fully immunized children, in Pentavalent vaccine coverage in infants and in tetanus toxoid vaccination in pregnant women (46–48). Conversely, a study in Vietnam, conducted in parallel to the present evaluation, found that while immunization data quality and use had significantly improved with the introduction of a paperless national immunization information system in two provinces, this was not associated with improvements in immunization outcomes, due to the interfering COVID-19 pandemic (49).

In both Guinea and Tanzania, use of an eLMIS enhanced reporting and tracking of vaccine doses. In Guinea, such a positive impact could be considered an indirect effect of the accompanying supportive supervision and quality assurance processes, given the lack of forecasting and ordering functionalities of the tool. In both countries, use of the tools was associated with better quality data on vaccine stock levels and fewer stockouts. Similarly, an earlier evaluation in Tanzania suggested that the eLMIS brought about performance improvements through better data use and management practices (50).

As described in the corresponding manuscript (29), eIR and eLMIS can potentially reduce costs and improve the efficiency of immunization data management and vaccine logistics in LMICs, but evidence across countries was mixed. The extent of cost savings was contingent on the degree to which the digital systems replaced traditional paper-based methods. The continued use of electronic tools could result in health workers being more efficient, organized, proactive, and able to immediately analyze and use the data generated. To achieve such an impact, systems will need to be used in real-time by first-line health workers at the service delivery level. To realize the full programmatic potential of electronic tools in LMICs there should be sufficient clarity on the strategic and programmatic goals of the use of eIR and eLMIS, the availability of the needed IT infrastructure and local IT expertise, the health service level of their implementation, and the level of integration into the HMIS and interoperability with the CRVS. Careful planning and investments are essential to realizing the full programmatic potential of electronic health tools in LMICs.

## Highlights

Four different electronic systems were evaluated in four LMICs, at different levels of their implementation, digitization and integration into the country HMIS.

- The implementation of eIR and eLMIS was associated with improved process efficiencies, specifically in service delivery, supervision and vaccine logistics.
- The noticeable digital literacy of health workers in LMICs provides confidence in the further digitization of health services.
- Electronic tools can be impactful if there is full government ownership and investments into a solid digital infrastructure.
- The migration to fully electronic systems will be essential to reap their benefits and to avoid paper-based duplications.
- Establishing interoperability of the eIR with CRVS will improve the ability of NIPs to reach unimmunized children.

## Authorship contribution statement

All authors attest they meet the ICMJE criteria for authorship. All authors contributed to all of the following: (1) the conception and design of the study, or acquisition of data, or analysis and interpretation of data, (2) drafting the article or revising it critically for important intellectual content, (3) final approval of the version to be submitted.

## Data Availability

All data produced in the present study are available upon reasonable request to the authors.

## Acknowledgements

The authors gratefully acknowledge the valuable contributions of the following colleagues to protocol design, field work, and data collection in the four countries: Alexandre Idrissa Baldè (EPI), François Loua (MOH), Alexandre Delamou (Africa Health) in Guinea; Gracia Odalys (PAHO), Marcela Contreras (PAHO), Gracia Velandia (PAHO), Leticia Puerto (MoH), Lourdes Otilia Mendoza (MoH), Flora Lopéz (MOH), Mariela Alvarado (MOH) in Honduras; Hassan Sibomana (RBC), Kizito Kayumba (MOH) in Rwanda; Florian Tinuga (MOH), Doreen Ibrahim Pamba (NIMR) in Tanzania; Liz Peloso and Chris Wright; and Claudio Jommi and Flaminia Sabbatucci (Bocconi University). The authors specifically thank the health workers, supervisors and caregivers of vaccinees for sharing their insights and perceptions on the use of the electronic tools at the service delivery level.

## Footnotes

1 EIR readiness assessment tool prepared by partners from US CDC, WHO HQ, PAHO, EURO, WPRO, UNICEF, BMGF, PATH, AIRA and supported by GAVI (reference pending)

2 https://www.kobotoolbox.org/

3 https://dhis2.org/about/

4 https://openlmis.org

5 http://openiz.org

6 https://immunizationdata.who.int/pages/coverage/DTP.html?CODE=GIN&ANTIGEN=DTPCV3&YEAR=

